# Longitudinal analysis of clinical serology assay performance and neutralising antibody levels in COVID19 convalescents

**DOI:** 10.1101/2020.08.05.20169128

**Authors:** Frauke Muecksch, Helen Wise, Becky Batchelor, Maria Squires, Elizabeth Semple, Claire Richardson, Jacqueline McGuire, Sarah Clearly, Elizabeth Furrie, Neil Greig, Gordon Hay, Kate Templeton, Julio C.C. Lorenzi, Theodora Hatziioannou, Sara Jenks, Paul D. Bieniasz

## Abstract

**Objectives:** To investigate longitudinal trajectory of SARS-CoV-2 neutralising antibodies and the performance of serological assays in diagnosing prior infection and predicting serum neutralisation titres with time

**Design:** Retrospective longitudinal analysis of a COVID19 case cohort.

**Setting:** NHS outpatient clinics

**Participants:** Individuals with RT-PCR diagnosed SARS-CoV-2 infection that did not require hospitalization

**Main outcome measures:** The sensitivity with which prior infection was detected and quantitative antibody titres were assessed using four SARS-CoV-2 serologic assay platforms. Two platforms employed SARS-CoV-2 spike (S) based antigens and two employed nucleocapsid (N) based antigens. Serum neutralising antibody titres were measured using a validated pseudotyped virus SARS-CoV-2 neutralisation assay. The ability of the serological assays to predict neutralisation titres at various times after PCR diagnosis was assessed.

**Results:** The three of the four serological assays had sensitivities of 95 to100% at 21-40 days post PCR-diagnosis, while a fourth assay had a lower sensitivity of 85%. The relative sensitivities of the assays changed with time and the sensitivity of one assay that had an initial sensitivity of >95% declined to 85% at 61-80 post PCR diagnosis, and to 71% at 81-100 days post diagnosis. Median antibody titres decreased in one serologic assay but were maintained over the observation period in other assays. The trajectories of median antibody titres measured in serologic assays over this time period were not dependent on whether the SARS-CoV-2 N or S proteins were used as antigen source. A broad range of SARS-CoV-2 neutralising titres were evident in individual sera, that decreased over time in the majority of participants; the median neutralisation titre in the cohort decreased by 45% over 4 weeks. Each of the serological assays gave quantitative measurements of antibody titres that correlated with SARS-CoV-2 neutralisation titres, but, the S-based serological assay measurements better predicted serum neutralisation potency. The strength of correlation between serologic assay results and neutralisation titres deteriorated with time and decreases in neutralisation titres in individual participants were not well predicted by changes in antibody titres measured using serologic assays.

**Conclusions:** SARS-CoV-2 serologic assays differed in their comparative diagnostic performance over time. Different assays are more or less well suited for surveillance of populations for prior infection versus prediction of serum neutralisation potency. Continued monitoring of declining neutralisation titres during extended follow up should facilitate the establishment of appropriate serologic correlates of protection against SARS-CoV-2 reinfection.

## Introduction

The emergence of severe acute respiratory syndrome coronavirus 2 (SARS-CoV-2), the causative agent of COVID-19, has resulted in a global pandemic with hundreds of thousands of deaths and millions of illnesses. Diagnosis of SARS-CoV-2 infection is principally dependent on RT-PCR using nasal and throat swabs, which is not ideally suited to mass population testing and as such has largely been targeted at symptomatic individuals in many settings. RT-PCR-diagnosed case numbers have therefore underestimated the prevalence of SARS-CoV-2 infection, and serology assays must be deployed to determine the true number of infections using a surveillance approach. Serology assays also have a critical role in screening volunteers for vaccine trials and convalescent plasma donations, as well as predicting infection or vaccine-induced immunity. Although several commercially available SARS-CoV-2 immunoassays are in common use, evaluation of their sensitivity has often used samples from hospitalised patients soon after infection. Knowledge of the long-term kinetics of antibody titres and the corresponding effectiveness of commercial assays is imperative if these testing protocols are to be accurately interpreted^1-3^.

Serology assays for SARS-CoV-2 primarily employ viral nucleocapsid (N) or the spike surface protein (S) antigens. Because S binds to target cells through its receptor binding domain (RBD) it is the target of neutralising antibodies. Therefore, S-based assays may be preferable to N-based assays for the assessment of the risk of future re-infection^4^. Of course, this premise is based on the assumptions (1) that neutralising antibodies constitute a major mechanism of protective immunity, and (2) that S-based serology assays accurately predict neutralising antibody activity.

Thus, major outstanding questions remain about the utilisation of serology that have implications for ongoing public heath testing strategies for SARS-CoV-2. These questions include (1) how circulating antibody levels that are specific for each viral antigen change with time following natural infection and (2) which serological assays best predict protective immunity. As yet, the prognostic value of antibody measurements in situations where individuals may be re-exposed to reinfection has yet to be demonstrated. Nevertheless, it is important to understand post infection serology as measured using high throughput assays to enable correlates of protection to be established. Here, we present the results of a longitudinal antibody testing study on a cohort of mildly symptomatic, non-hospitalised COVID19 positive patients during the first few months of convalescence. We compare the ability of four high-throughput automated assays to diagnose prior SARS-CoV2 infection and to predict neutralising activity in convalescent serum.

## Methods

### Participants

Participants with prior RT-PCR-diagnosed COVID-19 were recruited. Recruits were surveyed to determine the date of the positive PCR test, the date of onset of symptoms, and if their symptoms required hospitalisation. Serum samples were taken at a baseline visit (~3.5 to ~8.5 weeks post PCR test), and 2 weeks (visit 2), 4 weeks (visit 3) and 8 weeks later (visit 4). In total, 97 participants, who were not hospitalised during the course of their illness completed at least 3 visits. The mean age of the participants was 44.2 years (21 – 65 y), with 70 female (72% of cohort) participants. At visit 1 (baseline), the average number of days between PCR test and visit 1 (baseline) was 40.8 days (24 – 61 days); at visit 2 (2 weeks post-baseline), the average number of days post-PCR test was 55.1 days (40 – 79 days); at visit 3 (4 weeks post-baseline), the average number of days post-PCR test was 69.8 days (55 – 95 days); at visit 4 (8 weeks post-baseline), the average number of days post-PCR test was 98.4 days (85 – 110 days). Ethical approval was obtained for this study to be carried out through the NHS Lothian BioResource. All recruits gave written and informed consent for serial blood sample collection. Patients and Public were not involved in the design of this research.

### High throughput automated serology assays

Four commercial assays, that employ either S or N protein antigens and are designed for high throughput in healthcare settings were used. All the assays generate a qualitative positive/negative result based on assay-dependent signal thresholds. The Abbott SARS-CoV-2 IgG assay detects anti-N IgG using a two-step chemiluminescent microparticle immunoassay (CMIA) method with an acridinium-labelled anti-human IgG. The DiaSorin SARS-CoV-2 IgG assay is also a two-step CMIA method targeting undisclosed epitopes in the SARS-CoV-2 S protein and employs an isoluminol conjugated anti-human IgG. The Roche Anti-SARS-CoV total antibody assay is a two-step bridging electrochemiluminesent immunoassay (ECLIA) using ruthenium-labelled and biotin conjugated N protein. The Siemens SARS-CoV-2 total antibody assay is a one-step bridging CLIA method that detects antibodies against the RBD of the S protein, using acridinium and biotinylated S1 RBD. Assays were performed on the Abbott Architect and Diasorin Liason platforms (NHS Lothian), and the Roche Elecsys (NHS Lanarkshire) and Siemens Atellica (NHS Tayside) platforms. Serum, collected and stored according to the manufacturer’s recommendations, was used in all cases.

### SARS-CoV-2 Neutralisation assays

To measure neutralising antibody activity, serial dilutions of serum, beginning with a 1:12.5 dilution were five-fold serially diluted in 96-well plates over four dilutions. Thereafter, approximately 5×10^3^ infectious units of an HIV/CCNG/nLuc/SARS-CoV-2 pseudotype virus were mixed with the serum dilutions at a 1:1 ratio and incubated for 1 hour at 37 degrees in a 96-well plate. The mixture was then added to 293T/ACE2cl.22 target cells^5^ plated at 1×10^4^ cells/well in 100 ^l medium in 96-well plates the previous day. Thus, the final starting serum dilution was 1:50. Cells were cultured for 48h and harvested for NanoLuc luciferase assays, as previously described^5^.

## Results

The cohort studied herein consists of participants who were not hospitalised during the course of their illness and were therefore relatively mildly symptomatic. Participants were invited to report on the occurrence and frequency of symptoms. Approximately 70% of people reported at least one of the 3 main WHO -identified symptoms, namely fever, cough and anosmia. The most common of symptom was anosmia and the majority of participants reported the presence of 2 of these 3 symptoms (Table 1). Serum samples were collected from 97 participants at ~ 4 weeks (visit 1), 6 weeks (visit 2) and 8 weeks (visit 3) post diagnosis (by RT-PCR). Additionally, serum was collected from a subset (28 of the 97 participants) at ~ 12 weeks post diagnosis (visit 4).

**Table 1.** Percentage of participants per cohort displaying the three main WHO symptoms.

|  | **Fever** | **Cough** | **Anosmia** | **0 of 3 symptoms** | **1 of 3 symptoms** | **2 of 3 symptoms** | **All 3 symptoms** | **Self-reported recovered** |
| --- | --- | --- | --- | --- | --- | --- | --- | --- |
| Reported symptom | 65 | 69 | 74 | 1 | 19 | 42 | 35 | 44 |
| % | 67% | 71% | 76% | 1% | 16% | 43% | 36% | 49% |

N = 97 for all reported symptoms apart from "self-reported recovered", where only 90 individuals responded to this survey question

We compared the diagnostic sensitivity of 4 high throughput SARS-CoV-2 serology assays that are in routine use in hospital settings. Each assay gives a qualitative positive or negative result, based on assay specific thresholds and sensitivities were calculated for each assay using these thresholds. Inter and intra-assay analytical precision for each assay is detailed in Supplementary Table 1. To account for the differences in time post PCR diagnosis that participants made their first visit, sensitivity across a 20 day rolling time window was calculated. The Abbott, Roche and Siemens assays all had sensitivities of 95 to100% at 21-40 days post PCR-positive test, while the Diasorin assay had a lower sensitivity of 85% (fig 1A). However, the relative sensitivities of the assays changed with time. Specifically, the sensitivity of the Abbott assay declined to 85% in the 61-80 day window, and 71% at >81 days post diagnosis (fig 1A).

**Fig 1.**
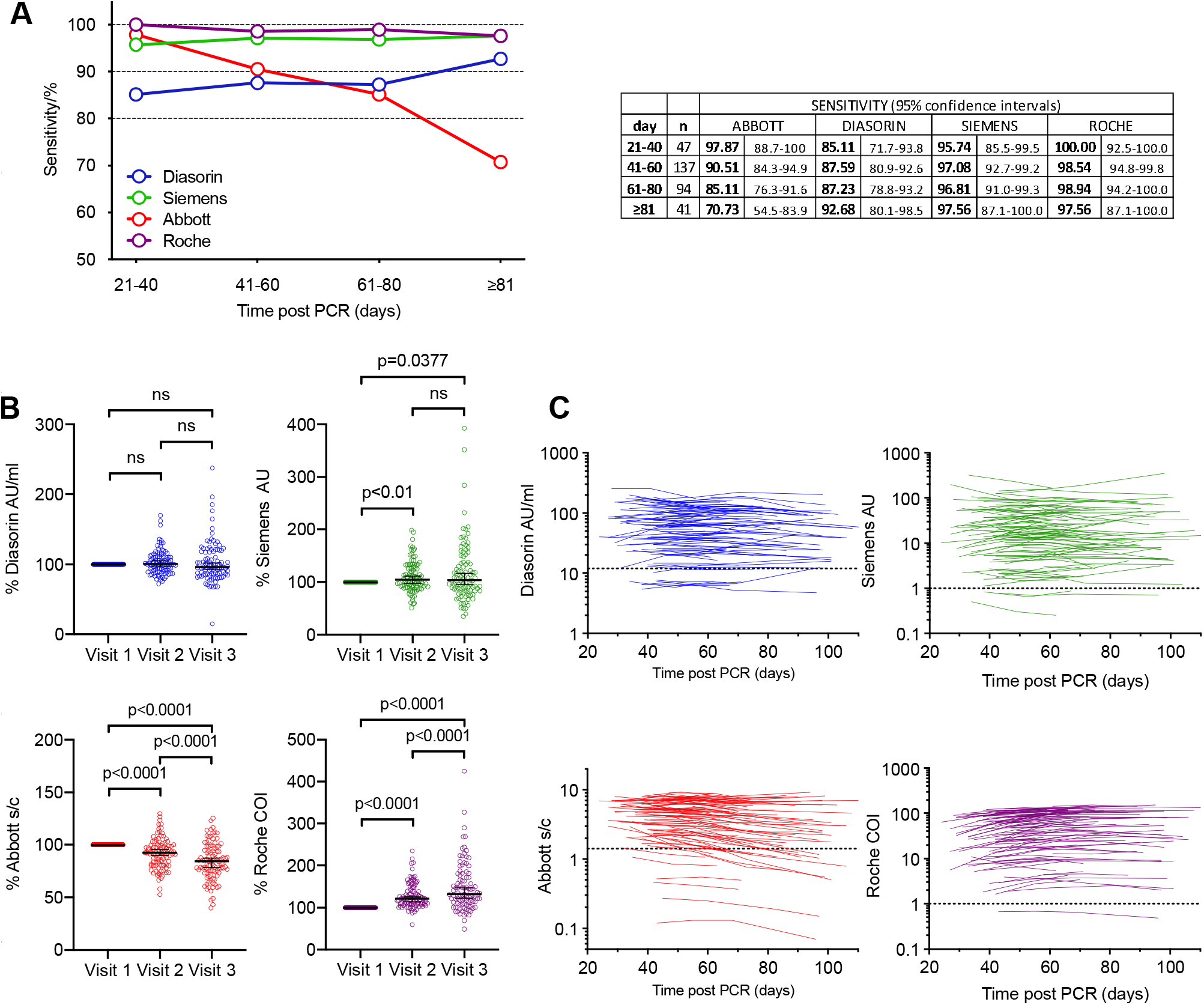
Longitudinal analysis of COVID-19 participant sera. **(A)** Sensitivity of the Abbott, Diasorin, Siemens and Roche serological assays (as indicated) measured in samples collected at four different timepoints, as indicated, post PCR test and 95% confidence interval. **(B)** Relative antibody titres for the Diasorin, Siemens, Abbott and Roche, assays at visits 1-3, normalized to visit 1. Horizontal line indicates median value with 95% confidence interval. Statistical significance was assessed with the Wilcoxon test. **(C)** Values forDiasorin, Siemens, Abbott and Roche serological assays for each participant plotted over time (each line represents one participant). Assay thresholds are indicated by a dotted horizontal line.

Conversely, the sensitivities of the other assays were maintained or increased over time (fig 1A). In terms of intra-individual change, 14/91 participants that were positive on the Abbott assay at visit 1 were negative by visit 3 or 4, whereas none of the participants with a positive result at visit 1 on the other assays became negative at visit 3 or 4. For the Diasorin assay, 2 participants that were negative at visit 1 were positive at visit 3 (both participants had an equivocal result at visit 1, and showed a small increase above the assay threshold at visit 3). In the Siemens assay, 3 participants were consistently negative, and in the Roche assay only a single participant was negative at each visit.

The serological assays give a quantitative assessment of antibody titre as well as a threshold-based positive/negative result. We next analysed changes in the quantitative results over time for each platform (fig 1 B, C). Mean antibody titres decreased in the Abbott assay at visits 2 and 3 compared to visit 1 (fig 1B) but increased in the Diasorin and particularly the Roche assays and remained approximately constant in the Siemens assay (Fig 1B). Notably, 79 out of 97 (81%) of participants showed a decrease in antibody titre on the Abbott platform, while 82/97 (85%) showed an increase on the Roche assay, despite the fact that both assays detect N-specific antibodies (fig 1 B, C). Negative or positive change was approximately equally likely in the S-based assays; specifically, 57% and 47% of intra-individual changes were negative for the Diasorin and Siemens assays respectively (fig 1 B, C).

We measured neutralising activity in serum samples from the first 3 visits for 80 of the 97 participants using a SARS-CoV-2 pseudotyped virus neutralisation assay. This assay employs HIV-1-based virions carrying a nanoluc luciferase reporter, pseudotyped with the SARS-CoV-2 spike protein. We have previously shown that neutralisation titres obtained using these pseudotyped particles correlate well with titres obtained using neutralisation of authentic SARS-CoV-2^5^, Moreover, this assay has been successfully applied for analysis of convalescent plasma samples and in a campaign to identify potent human monoclonal antibodies^6 7^. Consistent with our analyses of other cohorts^6 7^, a broad range of neutralising titres were evident in sera collected from 80 participants at three timepoints (fig 2A). In samples collected at visit 1, the neutralising activity, as determined by half-maximal neutralising titre (NT_50_), ranged from <30 to 4300, with a geometric mean of 234 (arithmetic mean was 411) (fig 2A, red symbols). Consistent with other cohorts ^6 7^ 34/80 (42%) had NT_50_ of less than 250 while only in 11/80 participants (14%) had NT_50_ values higher than 1000.

**Fig 2 -.**
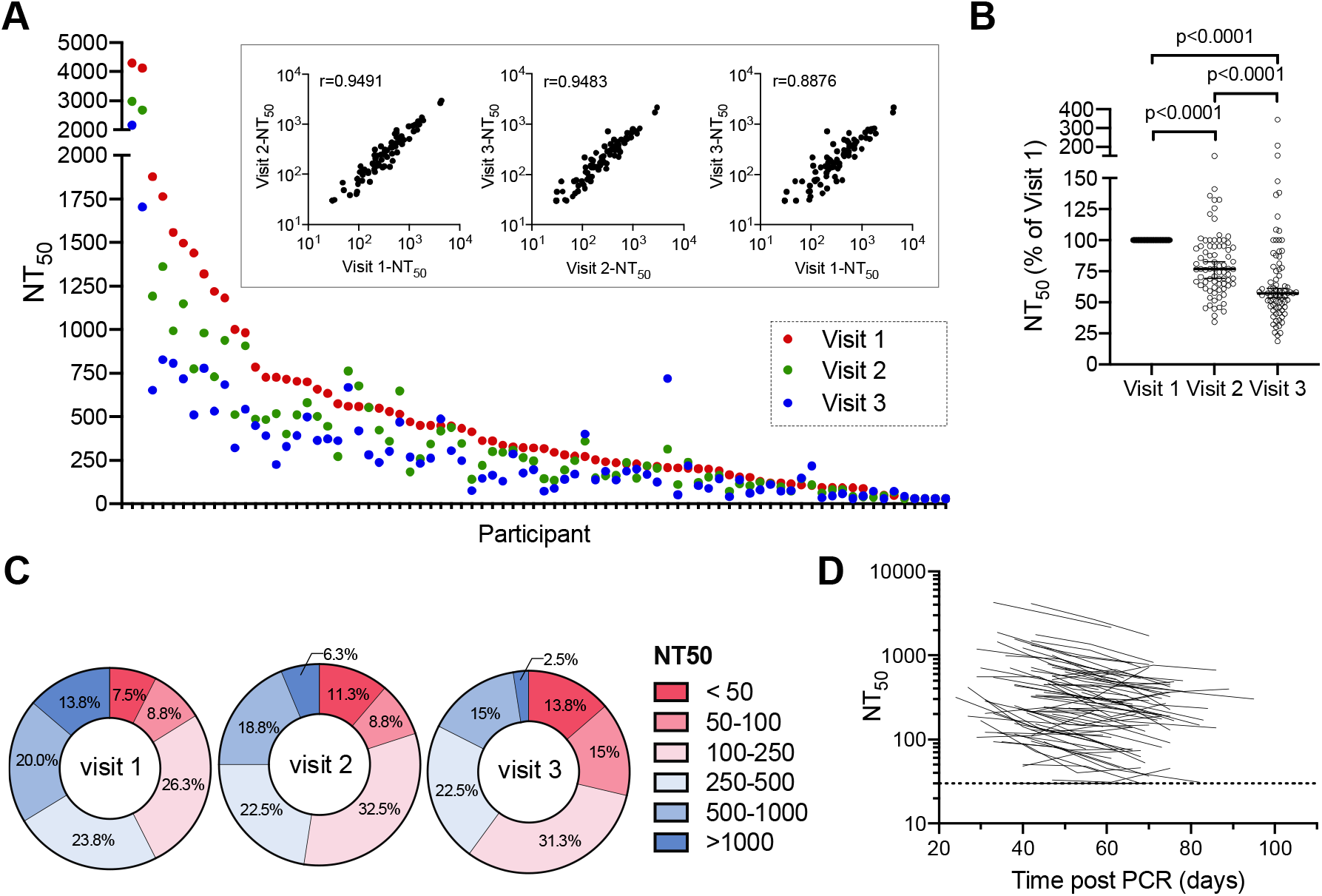
Neutralisation activity in COVID-19 participant sera. **(A)** Half-maximal neutralisation titres (NT_50_s) for each individual participant measured in serum samples collected at three different visits, as indicated by color. Inserts show correlation of NT_50_ values for samples collected at each visit, the spearman r is indicated (p<0.0001). **(B)** Relative NT_50_ values in sera obtained at visit 1 to 3, normalized to visit 1. Horizontal line represents median with 95% confidence interval. Statistical significance was assessed with the Wilcoxon test. **(C)** Frequency of sera with NT_50_ values falling to various quantitative categories at each visit. **(D)** NT_50_ values for each participant plotted over time (each line represents one participant). The limit of detection (LOD) is indicated by a dotted horizontal line.

NT_50_ values measured at each timepoint for individual participants correlated with each other, although there was divergence in NT_50_ values over time (fig 2 A inset). Notably, neutralising activity decreased at each time point for the majority of participants (fig 2 A, blue and green symbols). Overall, the decrease in median NT_50_ was ~25% per two-week sampling interval, resulting in a ~45% reduction in NT_50_ over the 4 weeks between visit 1 and visit 3 (fig 2B). As a result, distribution of NT_50_ values the cohort differed between visits (fig 2C). The relative decline in NT_50_ between visits 1 and 2 versus visits 2 and 3 did not differ significantly, and the majority of participants exhibited a similar relative decrease in neutralising activity over time, regardless of their initial NT_50_ values or the number of days post PCR at visit 1, suggesting exponential decay (fig 2D).

NT_50_ values at each sampling timepoint were poorly correlated with age (Supplementary fig 1A), and no correlation was observed between age and NT_50_ decay with time. As has been previously reported, there was a trend toward lower NT_50_ values in females than in males^6 7^, but there was no difference between sexes in NT_50_ decay with time (Supplementary fig 1B). Individual clinical parameters such as GI symptoms, fever or recovery time, did not predict NT_50_, serological values or decay parameters for any antibody measurement.

Next, we compared neutralising activity in serum with quantitative results obtained from the serological assays. Analysis of combined results from the three visits by 80 participants revealed a significant correlation between any combination of two serological assays (Supplementary fig 2).However, stronger correlations were observed between the two S-based assays, Siemens and Diasorin (r=0.92, p<0.0001) and between the two N-based assays Abbott and Roche (r=0.81, p<0.0001), The S-based assays correlated less well, but significantly (p<0.0001), with the N-based assays (Supplementary fig 2).

All the serological assays gave quantitative values that correlated with NT_50_ measurements, but as expected, the S-based assay measurements correlated more closely with NT_50_ measurements (fig 3A-D). The S1/S2-based Diasorin assay, was the best predictor of NT_50_ (r=0.84, p<0.0001, fig 3A), followed by the RBD-based Siemens assay (r=0.74, p<0.001, fig 3B), the N-based Abbott assay (r=0.69, p<0.0001, fig 3C) and, lastly, the Roche assay (r=0.56, p=0.0001, Fig 3D).

**Fig 3 -.**
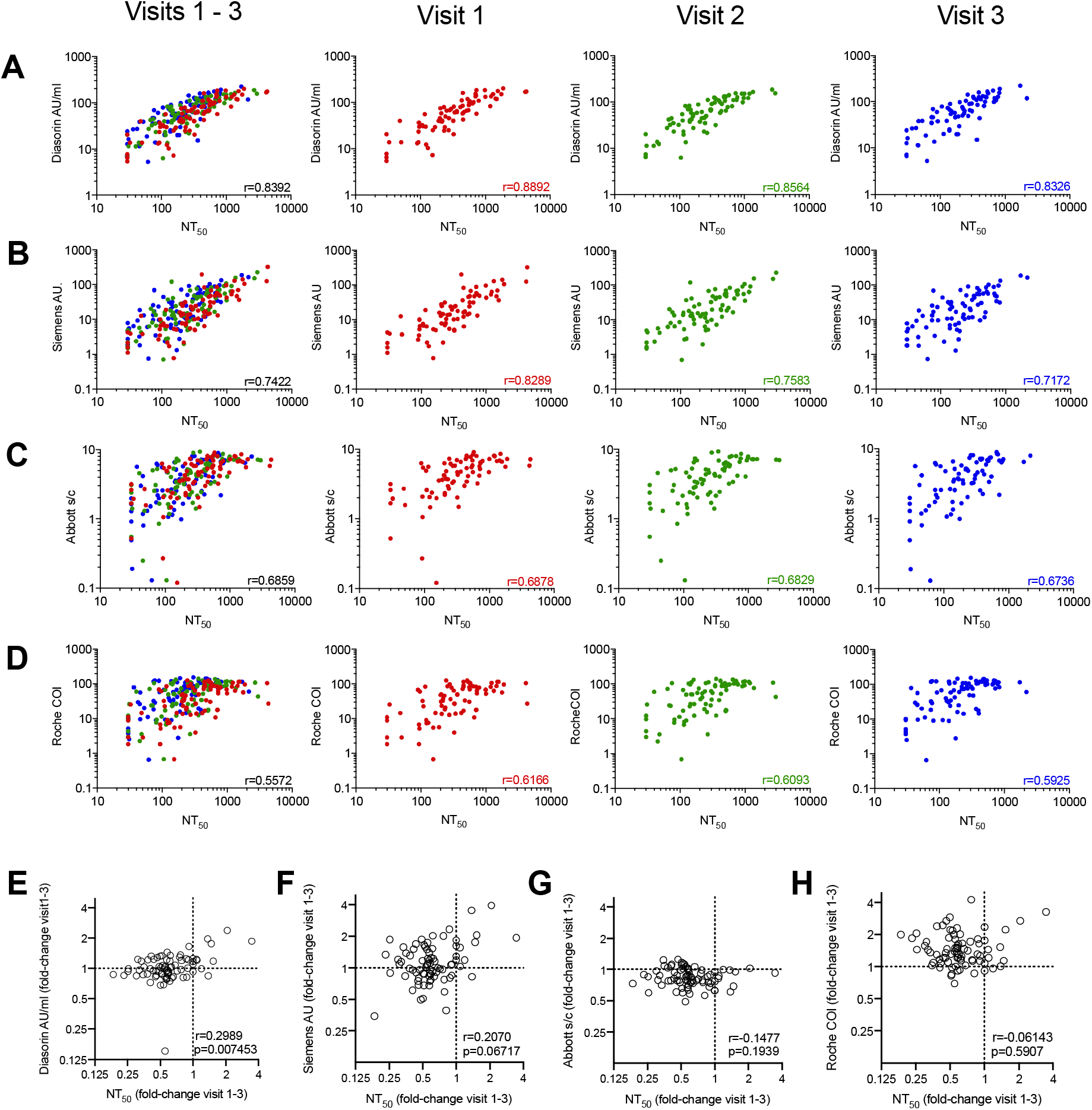
Correlation of serology results with neutralisation titres. **(A-D)** Serological assay values for the Diasorin **(A)**, Siemens **(B)**, Abbott **(C)** and Roche **(D)** assays versus NT_50_ values. Samples collected at each visit are indicated by color and are plotted individually as well as on a composite graph. Spearman r for all visits (black) and individual visits are indicated (p<0.0001). **(E-H)** Fold-change (visit 1 to visit 3) in serological assay values for the Diasorin **(E)**, Siemens **(F)**, Abbott **(G)** and Roche **(H)** assay versus fold-change in NT_50_ values. Spearman r and p-value are indicated.

The correlation between NT_50_ and the individual serological assays was best at the first visit and deteriorated to some extent thereafter (fig 3A-D, see color-coded r-values in individual graphs, p<0.0001 for all correlations), The decrease in the strength of correlation might, in part, be attributable to the fact that later sampling timepoints have more samples with lower NT_50_ values, which may reduce measurement precision. The magnitude of the deterioration in the predictive value differed between serological assays, with the S-based assays exhibiting larger decreases in correlation coefficients (r=0.89 and 0.83 at visit 1, versus r=0.83 and 0.71 at visit 3 for DiaSorin and Siemens assays respectively fig 3A-D), Despite the increasing disparity over time, the DiaSorin assay was clearly superior at predicting NT_50_ at all visits (fig 3A-D).

Interestingly, comparison of the extent of change in neutralisation activity over the 4-week observation interval with the concomitant change in values obtained using serological assays, revealed only minimal correlation (fig 3E-H, supplementary fig 3). Notably, in most participants, the decline in serum neutralising activity was clearly greater than the decline in antibody titre measured using any serological assay (fig 3E-H supplementary fig 3). Even for the Diasorin assay, which gave the best prediction of neutralising activity at each time point (fig 3A), declines in neutralising activity were not well predicted by declines in Diasorin assay measurements (fig 3E, supplementary fig 3). While both the Abbott assay and the NT_50_ measurements exhibited declining antibody titres with time, the magnitudes of these declines did not correlate with each other (fig 3G, supplementary fig 3).

## Discussion

Serological assays for infectious agents have two major and distinct uses, namely (1) to diagnose chronic infections (e.g. HIV-1) and (2) to determine past infection or immunisation status (e.g. measles, VZV) which may be able to predict immunity from future infection. For example, HIV-1 and HCV serological tests are crucial for diagnosis but have no role in prediction of immunity. In contrast, for viruses such as VZV and measles, the main role of serology is to predict immunity induced by vaccination or prior infection. The use of serological assays to determine widespread seroprevalence is a relatively new application following the COVID19 pandemic, which is different from how these assays have traditionally been used clinically and requires understanding of how these assays perform in populations over time.

During the current SARS-CoV-2 pandemic it has become clear that the magnitude of serologic immune responses is highly variable^6 7^. Nevertheless, the vast majority of individuals with a PCR-confirmed SARS-CoV-2 infection generate antibodies at a sufficient level for diagnosis of recent infection^8^. A number of commercial assays have been deployed for high throughput SARS-CoV-2 antibody testing in a clinical setting, and evaluated mostly using hospitalized participants^9 10^. Non-hospitalised patients and those with mild disease typically have lower levels of antibodies than hospitalized patients with severe illness^11-15^, Using our cohort of non-hospitalized participants with mild disease, all four assays evaluated herein had sensitivities at visit 1 (an average of 40.8 days after PCR testing) that were comparable to the evaluations performed for these platforms using hospitalised patients^16^. This would therefore make all four assays suitable for the detection of COVID-19 antibodies shortly after infection as a confirmatory test for diagnostic purposes, when used in conjunction with RT-PCR assays and clinical history

However, differences in assay diagnostic sensitivity become apparent at later time points. Specifically, the sensitivity of the widely used Abbott assay declined with time, to ~70% at >81 days post PCR. Consequently, this assay is not appropriate for seroprevalence studies, for identification of SARS-CoV-2 naive vaccine trial participants, or for investigation of individuals presenting with long term chronic symptoms. Altering the positive/negative threshold, may mitigate this issue^17^, but would not ultimately alter the downward trend in assay signal over time. Notably our study is one of the few that would capture this information, as most other studies have examined seroconversion at early time points^14 18-20^. Reasons for the differences in assay performance over time are unclear but cannot be attributed solely to the choice of antigen. Although other studies have attributed a decline in sensitivity of an N based assay to an inherent difference in the dynamics of S versus N antibodies^21^ our findings do not support this contention, at least during the first ~100 days of convalescence. Both Abbott and Roche assays employ the N-proteins as an antigen, but Abbott assay titres decline while those in the Roche assay increase during this time period. One possible explanation for this difference is the use of an antigen bridging approach in the Roche assay, where declines in the total amount of antibody might be compensated by increases in affinity or avidity as antibodies mature through somatic hypermutation. Alternatively, it is possible that the range of N epitopes recognized by sera might change with time. Whatever the explanation, it is clear that that the trajectories of antibody titres measured using assays based on recognition of the same or related antigens can differ^22-25^.

Overall, given their superior sensitivity at each of the time points investigated thus far, the Siemens and Roche assays appear most appropriate for diagnosis of prior SARS-CoV-2 infection, at least within 4 months of SARS-CoV-2 infection, and would report a higher population prevalence than Abbott or DiaSorin assays in the 1 to 4 month post infection period.

While the Roche assay exhibited the best diagnostic sensitivity and is therefore well suited for serosurveillance during this time period, it had the lowest ability to predict neutralising antibody titres. This finding might be expected, as neutralising antibodies are directed to the S protein while N-specific antibodies are not expected to be neutralising. The Diasorin assay best predicted neutralising titres, and marginally outperformed the Siemens assay in this regard, perhaps because the dominant neutralising and/or S-binding activity in at least some sera is provided by antibodies that recognize epitopes outside the RBD^26 27^. It is important to recognize however, that many S-binding antibodies are not neutralising - measurements of S-binding antibodies remain correlates of, and not direct measures of, neutralising antibodies^7^.

Very recent reports have also indicated that neutralising antibody titres decline with time^23^ ^24^, while another study reported that neutralisation titres remained stable for at least 3 months post infection^28^. However, in the latter case neutralisation titres were inferred based on a serologic ELISA measurement that was calibrated using a neutralisation assay performed on a small subset of samples. As shown herein, neutralising antibody levels indeed decline in most patients, even those who apparently maintain S-binding or RBD-binding antibody titres measured in serology assays. Thus, the trajectory of neutralising antibody levels cannot necessarily be deduced from serologic measurements of S-binding antibodies, even though S-binding antibodies and neutralising titres are broadly correlated.

Key questions in SARS-CoV-2 serology are the trajectory of of the antibody response and to what degree the titres of neutralising antibodies, or antibodies that simply bind to S or N correlate with protection from reinfection or severe disease. Serological studies based on hCoV infection have shown that many adults possess detectable circulating antibodies against OC43 and 229E^29^, and children seroconvert to NL63 and 229E before ~3.5 years of age^30^. These baseline levels increased upon infection, returning to baseline within one year. High levels of circulating neutralising antibody correlated with protection from re-infection with the same strain of virus^31 32^. However, hCoV re-infections occur^32 33^, with more mild illness and shorter duration of virus shedding. Thus, in the case of seasonal hCoV, these data suggest that immunity may wane over time. More limited data is available for SARS-CoV and MERS-CoV, although it suggests antibody responses also decline in the majority of infected individuals^34^.

If, as seems plausible, neutralising antibodies constitute a major protective mechanism against SARS-CoV-2 infection, then the use of serological assays that use S-based antigens and correlate best with NT_50_ measurements would appear most appropriate for prognostication of immunity. Conversely, if other mechanisms of immunity, such as long-lived memory T-cell responses play a dominant role in protection from infection or severe disease^35-38^, then the optimal choice of antigen for serology assays might differ. Because detailed analyses of T-cell responses are not currently feasible in a high throughput clinical setting, future work should examine the frequency of reinfection and clinical outcomes in cohorts with detailed longitudinal analyses of serum antibodies to both N and S antigens to determine the prognostic value of such measurements.

## Data Availability

Raw de-identified data for serology measurements and NT50 measurements is available from the authors on request.

## Acknowledgements

We acknowledge the support and hard work of the NHS Lothian BioResouce team including in particular David Harrison for his support with initiating this study; Frances Rae and Craig Marshall for their advice and Linda MacLeod for her advice and hard work. NHS Lothian laboratories and out-patients teams have also worked extremely hard to enable the provision of this research study. In particular we would like to thank Joan Donnelly for her support and Susan Taylor for her dedication and hardwork.

## Contributor and guarantor information

Contributors: HW, SJ, TH and PDB conceived and designed the study. HW, BB, MS, ES, CR, JM, SC, EF, NG, GH, KT and SJ acquired and analysed data using the serological assay platforms. FM and JCCL performed the neutralisation assays. FM and TH did additional data analysis. HM, BB, SJ and PDB wrote the first draft of the manuscript. HM, BB, FM, TH, SJ and PB critically reviewed and revised the draft. All authors approved the final version of the manuscript for submission. HW, BB and MS contributed equally. SJ and PDB are the guarantors. The corresponding author attests that all listed authors meet authorship criteria and that no others meeting the criteria have been omitted.

## Copyright statement

The Corresponding Author has the right to grant on behalf of all authors and does grant on behalf of all authors, an exclusive licence on a worldwide basis to the BMJ Publishing Group Ltd to permit this article (if accepted) to be published in BMJ editions and any other BMJPGL products and sublicences such use and exploit all subsidiary rights, as set out in our licence

## Competing interest statement

All authors have completed the Unified Competing Interest form (available on request from the corresponding author) and declare: no support from any organisation or financial relationships with any organisations that might have an interest in the submitted work in the previous three years, no other relationships or activities that could appear to have influenced the submitted work.

## Transparency declaration

Paul Bieniasz and Sara Jenks (Guarantors) affirm that the manuscript is an honest, accurate, and transparent account of the study being reported; that no important aspects of the study have been omitted; there were no discrepancies from the study as planned.

## Details of ethical approval

Ethical approval was obtained for this study to be carried out through the NHS Lothian BioResource. All recruits gave written and informed consent for serial blood sample collection. The Rockefeller University IRB reviewed and approved the study.

## Funding

This work was supported by the NHS and Grants from the National Institutes of Allergy and infectious Diseases R37AI640003 (to PDB) and R01AI078788 (to TH). There were no study sponsors. The funders played no role in the design, analysis or reporting of this research.

## Data sharing statement

Raw de-identified data for serology measurements and NT_50_ measurements is available from the authors on request. we plan to disseminate the results to study participants and or patient organisations

## Checklist

A filled in checklist for appropriate study type is attached

## Supplementary table

**Supplementary Table 1.** Inter-assay and intra-assay precision for main analyser serological assays.

|  |  | Intra-assay precision<sup>1</sup> |  |  | Inter-assay precision<sup>1</sup> |  |  |
| --- | --- | --- | --- | --- | --- | --- | --- |
|  |  | Mean | SD | CV (%) | Mean | SD | CV (%) |
| Roche Cobas e 801 COI | Negative | 0.09 | 0.0025 | 2.91 | 0.09 | 0.006 | 7.54 |
|  | Positive | 13.85 | 0.14 | 1.00 | 13.85 | 0.44 | 3.14 |
| Abbott Architect i2000 S/C | Negative | 0.03 | 0.001 | 2.94 | 0.03 | 0.002 | 6.57 |
|  | Positive | 8.11 | 0.04 | 0.50 | 8.11 | 0.07 | 1.20 |
| Siemens Atellica AU | Negative | 0.05 | 0.0039 | 7.70 | 0.05 | 0.009 | 1.70 |
|  | Positive | 2.82 | 0.05 | 1.80 | 2.82 | 0.10 | 3.70 |
| DiaSorin LIASON AU/ml | Negative | 6.01 | 0.20 | 3.40 | 6.13 | 0.30 | 4.83 |
|  | Positive | 69.72 | 1.06 | 1.52 | 69.10 | 1.98 | 2.86 |

<sup>1</sup>Based on negative and positive pooled patient material. Inter-assay precision was determined using the mean of at least 5 replicates run on 3- 5 separate days for each assay.

## Supplementary Figures

**Supplementary fig 1.**
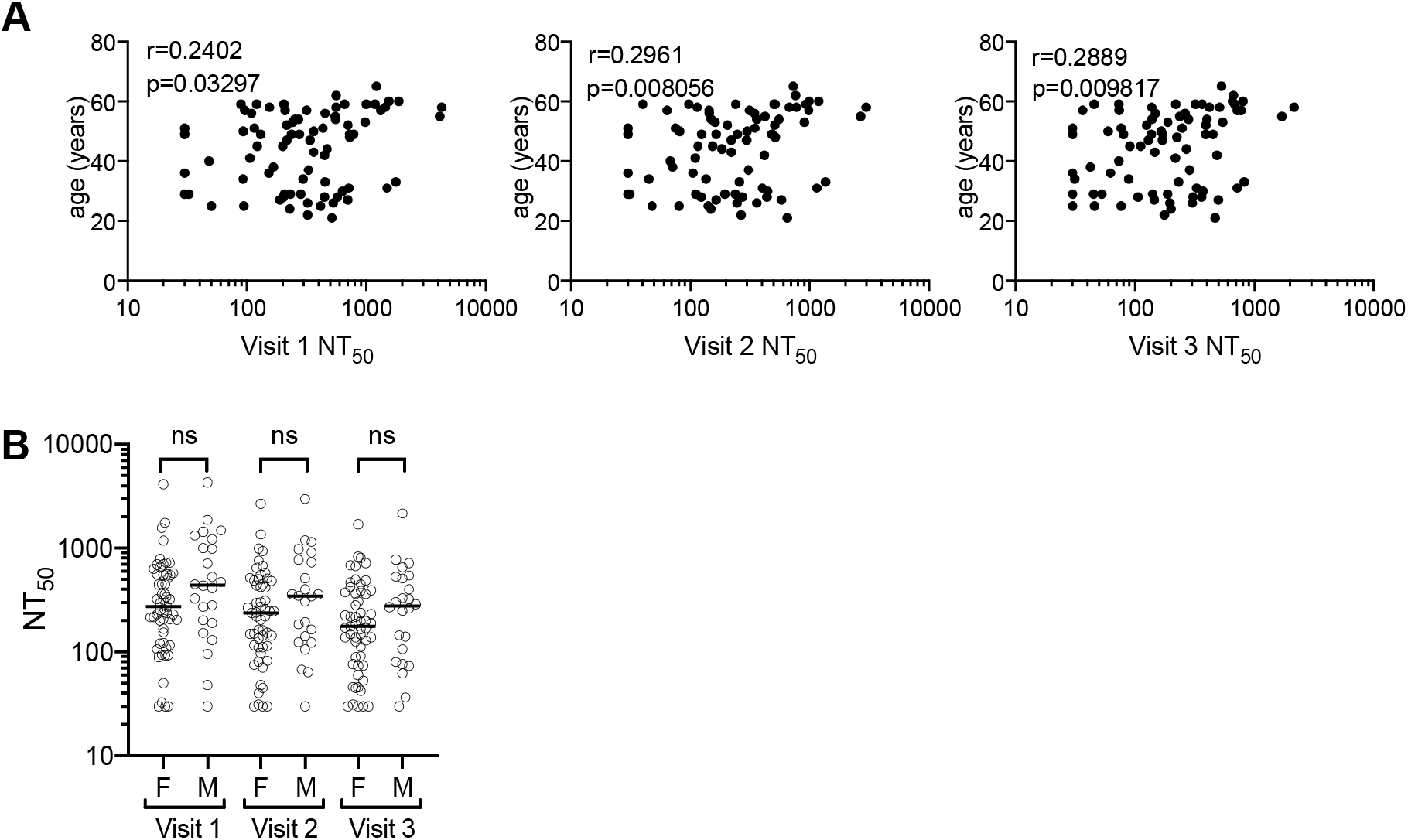
**(A)** Age (years) versus NT_50_ measured in sera collected at each visit. Spearman r and respective p-values are indicated. **(B)** NT_50_ in sera from male and female participants, collected at visits 1, 2 and 3. Horizontal lines indicate median values. Statistical significance was assessed with the Mann-Whitney-test.

**Supplementary fig 2.**
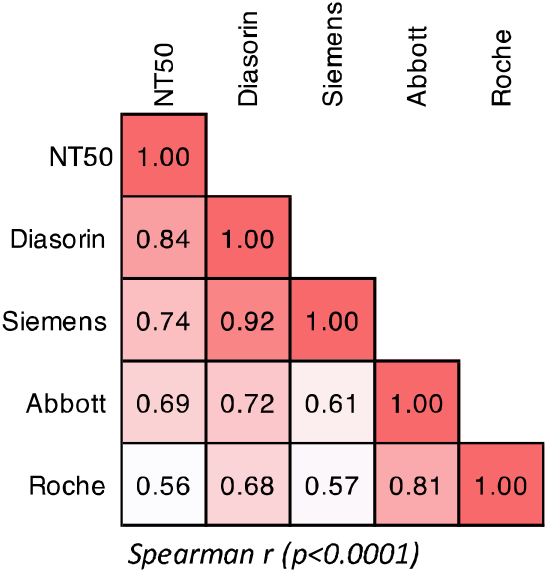
**(A)** Spearman r for correlations of serological assay measurements and NT_50_ for all samples analysed (not divided by vist).

**Supplementary fig 3.**
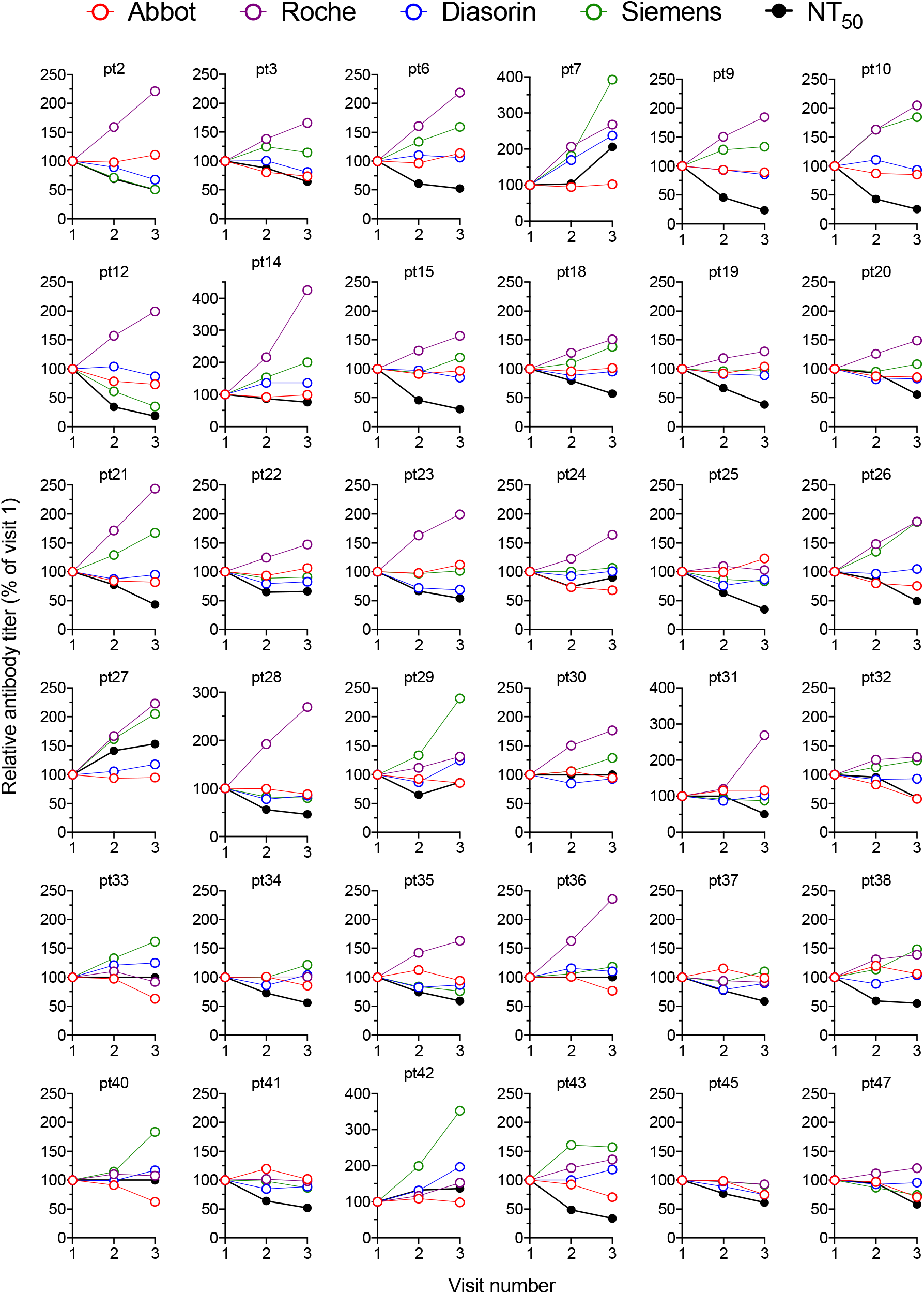
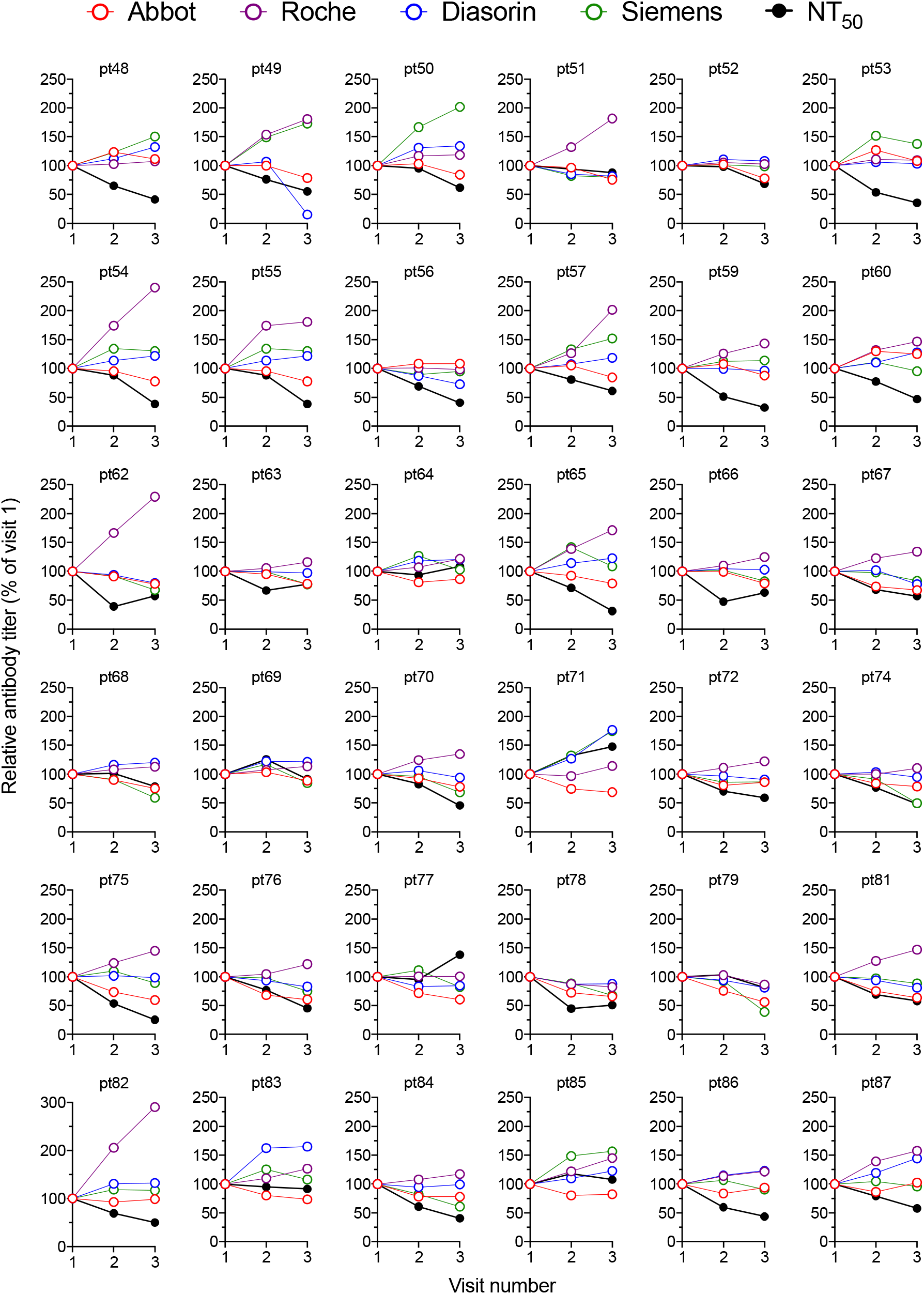
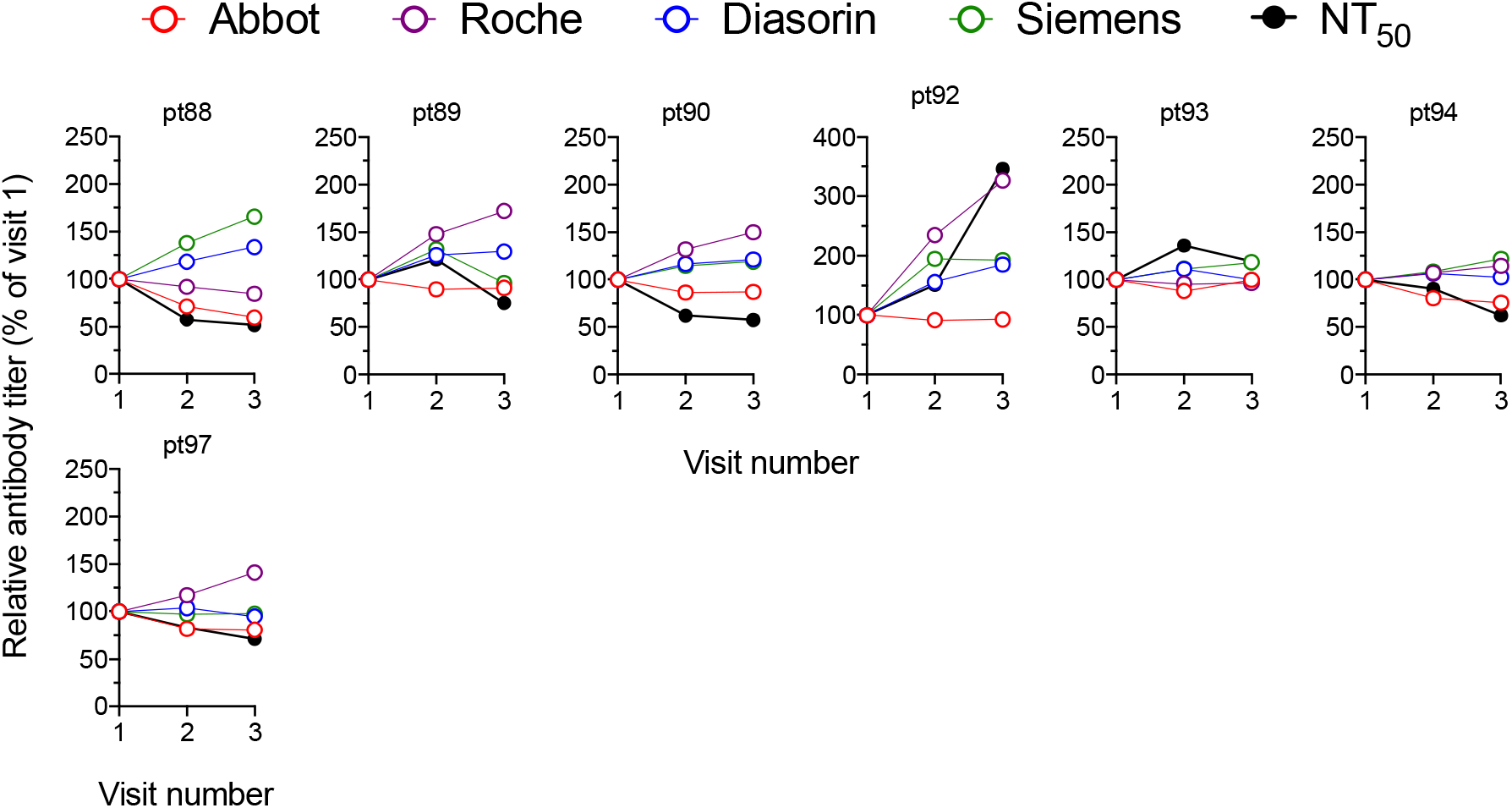
Changes in antibody titres measured using each serology platfrom and using the neutralization assay (relative to visit one which is plotted as 100%) at each visit for each individual participant.

## Notes

### Competing Interest Statement

The authors have declared no competing interest.

### Author Declarations

NHS Lothian BioResource

## References

1. Andersson M, Low N, French N, et al. Rapid roll out of SARS-CoV-2 antibody testing—a concern. BMJ 2020:m2420. doi: 10.1136/bmj.m2420

2. Özçürümez MK, Ambrosch A, Frey O, et al. SARS-CoV-2 antibody testing—questions to be asked. Journal of Allergy and Clinical Immunology 2020;146(1):35–43. doi: 10.1016/j.jaci.2020.05.020

3. GeurtsvanKessel CH, Okba NMA, Igloi Z, et al. An evaluation of COVID-19 serological assays informs future diagnostics and exposure assessment. Nature Communications 2020;11(1):3436. doi: 10.1038/s41467-020-17317-y

4. Hess AS, Shardell M, Johnson JK, et al. Methods and recommendations for evaluating and reporting a new diagnostic test. European Journal of Clinical Microbiology & Infectious Diseases 2012;31(9):2111-16. doi: 10.1007/s10096-012-1602-1

5. Schmidt F, Weisblum Y, Muecksch F, et al. Measuring SARS-CoV-2 neutralizing antibody activity using pseudotyped and chimeric viruses. Journal of Experimental Medicine 2020;217(11):e20201181. doi: 10.1084/jem.20201181

6. Luchsinger LL, Ransegnola B, Jin D, et al. Serological Analysis of New York City COVID19 Convalescent Plasma Donors: Infectious Diseases (except HIV/AIDS), 2020.

7. Robbiani DF, Gaebler C, Muecksch F, et al. Convergent antibody responses to SARS-CoV-2 in convalescent individuals. Nature 2020 doi: 10.1038/s41586-020-2456-9

8. Wajnberg A, Mansour M, Leven E, et al. Humoral immune response and prolonged PCR positivity in a cohort of 1343 SARS-CoV 2 patients in the New York City region: Infectious Diseases (except HIV/AIDS), 2020.

9. Group TNS-C-SAEGTNS-C-SAE. Head-to-head benchmark evaluation of the sensitivity and specificity of five immunoassays for SARS-CoV-2 serology on <1500 samples. 2020 doi: 10.6084/M9.FIGSHARE.C.5046032.V1

10. Jääskeläinen A, Kuivanen S, Kekäläinen E, et al. Performance of six SARS-CoV-2 immunoassays in comparison with microneutralisation. Journal of Clinical Virology 2020;129:104512. doi: 10.1016/j.jcv.2020.104512

11. Cervia C, Nilsson J, Zurbuchen Y, et al. Systemic and mucosal antibody secretion specific to SARS-CoV-2 during mild versus severe COVID-19: Immunology, 2020.

12. Dogan M, Kozhaya L, Placek L, et al. Novel SARS-CoV-2 specific antibody and neutralization assays reveal wide range of humoral immune response during COVID-19: Allergy and Immunology, 2020.

13. Klein S, Pekosz A, Park H-S, et al. Sex, age, and hospitalization drive antibody responses in a COVID-19 convalescent plasma donor population: Infectious Diseases (except HIV/AIDS), 2020.

14. Long Q-X, Liu B-Z, Deng H-J, et al. Antibody responses to SARS-CoV-2 in patients with COVID-19. Nature Medicine 2020 doi: 10.1038/s41591-020-0897-1

15. Rijkers G, Murk J-L, Wintermans B, et al. Differences in antibody kinetics and functionality between severe and mild SARS-CoV-2 infections.: Infectious Diseases (except HIV/AIDS), 2020.

16. Perkmann T, Perkmann-Nagele N, Breyer M-K, et al. Side by side comparison of three fully automated SARS-CoV-2 antibody assays with a focus on specificity: Infectious Diseases (except HIV/AIDS), 2020.

17. Bryan A, Pepper G, Wener MH, et al. Performance Characteristics of the Abbott Architect SARS-CoV-2 IgG Assay and Seroprevalence in Boise, Idaho. J Clin Microbiol 2020;58(8) doi: 10.1128/jcm.00941-20 [published Online First: 2020/05/10]

18. Lou B, Li T-D, Zheng S-F, et al. Serology characteristics of SARS-CoV-2 infection since exposure and post symptom onset. European Respiratory Journal 2020:2000763. doi: 10.1183/13993003.00763-2020

19. Pickering S, Betancor G, Pedro Galao R, et al. Comparative assessment of multiple COVID-19 serological technologies supports continued evaluation of point-of-care lateral flow assays in hospital and community healthcare settings: Infectious Diseases (except HIV/AIDS), 2020.

20. Staines HM, Kirwan DE, Clark DJ, et al. Dynamics of IgG seroconversion and pathophysiology of COVID-19 infections: Infectious Diseases (except HIV/AIDS), 2020.

21. Grandjean L, Saso A, Ortiz A, et al. Humoral Response Dynamics Following Infection with SARS-CoV-2. *medRxiv* 2020

22. Perreault J, Tremblay T, Fournier M-J, et al. Longitudinal analysis of the humoral response to SARS-CoV-2 spike RBD in convalescent plasma donors: Immunology, 2020.

23. Ibarrondo FJ, Fulcher JA, Goodman-Meza D, et al. Rapid Decay of Anti-SARS-CoV-2 Antibodies in Persons with Mild Covid-19. New England Journal of Medicine 2020:NEJMc2025179. doi: 10.1056/NEJMc2025179

24. Seow J, Graham C, Merrick B, et al. Longitudinal evaluation and decline of antibody responses in SARS-CoV-2 infection: Infectious Diseases (except HIV/AIDS), 2020.

25. Juno JA, Tan H-X, Lee WS, et al. Humoral and circulating follicular helper T cell responses in recovered patients with COVID-19. Nature Medicine 2020 doi: 10.1038/s41591-020-0995-0

26. Weisblum Y, Schmidt F, Zhang F, et al. Escape from neutralizing antibodies by SARS-CoV-2 spike protein variants: Microbiology, 2020.

27. Barnes CO, West AP, Huey-Tubman KE, et al. Structures of human antibodies bound to SARS-CoV-2 spike reveal common epitopes and recurrent features of antibodies. Cell 2020:S0092867420307571. doi: 10.1016/j.cell.2020.06.025

28. Wajnberg A, Amanat F, Firpo A, et al. SARS-CoV-2 infection induces robust, neutralizing antibody responses that are stable for at least three months: Infectious Diseases (except HIV/AIDS), 2020.

29. Macnaughton MR. Occurrence and frequency of coronavirus infections in humans as determined by enzyme-linked immunosorbent assay. Infection and Immunity 1982;38(2):419–23. doi: 10.1128/IAI.38.2.419-423.1982

30. Dijkman R, Jebbink MF, Gaunt E, et al. The dominance of human coronavirus OC43 and NL63 infections in infants. Journal of Clinical Virology 2012;53(2):135–39. doi: 10.1016/j.jcv.2011.11.011

31. Callow KA. Effect of specific humoral immunity and some non-specific factors on resistance of volunteers to respiratory coronavirus infection. The Journal of Hygiene 1985;95(1):173–89. doi: 10.1017/s0022172400062410

32. Callow KA, Parry HF, Sergeant M, et al. The time course of the immune response to experimental coronavirus infection of man. Epidemiology and Infection 1990;105(2):435–46. doi: 10.1017/S0950268800048019

33. Edridge AW, Kaczorowska JM, Hoste AC, et al. Coronavirus protective immunity is short-lasting: Infectious Diseases (except HIV/AIDS), 2020.

34. Kellam P, Barclay W. The dynamics of humoral immune responses following SARS-CoV-2 infection and the potential for reinfection. Journal of General Virology 2020 doi: 10.1099/jgv.0.001439

35. Gallais F, Velay A, Wendling M-J, et al. Intrafamilial Exposure to SARS-CoV-2 Induces Cellular Immune Response without Seroconversion: Infectious Diseases (except HIV/AIDS), 2020.

36. Grifoni A, Weiskopf D, Ramirez SI, et al. Targets of T cell responses to SARS-CoV-2 coronavirus in humans with COVID-19 disease and unexposed individuals. Cell 2020:S0092867420306103. doi: 10.1016/j.cell.2020.05.015

37. Le Bert N, Tan AT, Kunasegaran K, et al. SARS-CoV-2-specific T cell immunity in cases of COVID-19 and SARS, and uninfected controls. Nature 2020 doi: 10.1038/s41586-020-2550-z

38. Sekine T, Perez-Potti A, Rivera-Ballesteros O, et al. Robust T cell immunity in convalescent individuals with asymptomatic or mild COVID-19: Immunology, 2020.

